# Charity Care, Uncompensated Care, and Medicaid Patient Volume Among 340B and Non-340B Hospitals

**DOI:** 10.64898/2026.02.12.26346191

**Authors:** Robert Popovian, Anne M. Sydor, Kim Czubaruk, Michael Walker, William Smith

## Abstract

**Background:** The 340B Drug Pricing Program allows eligible safety-net hospitals and clinics to purchase outpatient drugs from manufacturers at discounted prices, creating revenue that can support care for low-income and uninsured patients. As the program has expanded, questions have emerged about whether participating hospitals’ reported financial assistance and patient-volume measures align with the program’s intended objectives.

**Objectives:** We compared 340B and non-340B hospitals using reported measures related to care for uninsured and underserved patients, including charity care, charity care for uninsured patients, uncompensated care, and Medicaid admission days.

**Methods:** We conducted a cross-sectional analysis comparing unadjusted mean values across 340B and non-340B hospital categories. 340B designation was derived by linking hospitals in RAND-transposed CMS HCRIS hospital cost report data to HRSA 340B covered-entity records. Hospitals identified as active 340B covered entities during the relevant reporting period were classified as 340B participants; all others were classified as non-340B. Key measures included charity care as a percentage of operating expenses, Medicaid admission days as a share of hospital days, unreimbursed or uncompensated care, and costs associated with uninsured patients approved for charity care. Welch’s t-tests assessed statistically significant differences in unadjusted mean values.

**Results:** Among 3,999 hospitals, 340B hospitals reported lower charity care than non-340B hospitals (2.16% vs 2.82% of operating expenses, P<.05) and lower charity care for uninsured patients (1.60% vs 2.26%, P<.05). However, 340B hospitals reported higher Medicaid admission days (19.69% vs 17.76%, P<.05). Unreimbursed or uncompensated care was numerically lower among 340B hospitals, but the overall difference was not statistically significant.

**Conclusion:** These findings raise accountability questions about whether 340B participation consistently corresponds with greater reported direct financial assistance for uninsured and underserved patients. Greater transparency, standardized reporting, and clearer patient-assistance expectations may help ensure that 340B program benefits are better aligned with the needs of vulnerable populations.

## Introduction

The 340B Drug Pricing Program, enacted in 1992, is a foundational component of the U.S. safety-net infrastructure, enabling qualifying hospitals and clinics, known as covered entities, to procure outpatient medicines at significantly discounted prices from biopharmaceutical manufacturers, with such discounts subsequently being arbitraged by the 340B hospital to generate revenue.^1^ The program was originally intended to allow eligible safety-net providers, including disproportionate share hospitals (DSH), to stretch limited resources, reach more eligible patients, and provide more comprehensive care to low-income and uninsured populations.^2^

Over subsequent decades, covered entities have substantially benefited from these manufacturer discounts, which they have used to stabilize finances and expand services. However, as the 340B program has grown in both scale and complexity, it has evolved in ways that have prompted critical examination.^3^

### Expansion under the ACA and Proliferation of Contract Pharmacies

The Patient Protection and Affordable Care Act (ACA) expanded eligibility for the 340B program to include rural referral centers (RRC), sole community hospitals (SCH), critical access hospitals (CAH), and certain free-standing children’s and cancer hospitals.^4^ This expansion significantly increased the number of covered entities in the program.^5,6^

In 2010, following the passage of the ACA, the Health Resources and Services Administration (HRSA) issued guidance permitting covered entities that lacked an in-house pharmacy to contract with multiple external pharmacies rather than a single pharmacy.^7^ This regulatory shift led to a dramatic increase in the number of contract pharmacies across the United States.^6^

### Declining Charity Care, Revenue Accumulation, and Transparency Concerns

While the 340B program has expanded, research by the Pioneer Institute, using RAND-transposed Centers for Medicare & Medicaid Services Healthcare Cost Report Information System (CMS HCRIS) hospital cost report data, indicates that hospital charity care levels have decreased, raising concerns about whether the program’s benefits are being used as intended.^6^

This trend has led to calls for greater transparency, including requirements that hospitals disclose the amount of revenue derived from 340B discounts and the amount reinvested in charity care.

Currently, 340B-eligible hospitals are not required to report their 340B-related revenue or how it is allocated publicly.^8^

Some states, however, have taken legislative action. Minnesota, for example, has enacted a law mandating that hospitals disclose detailed financial data, including 340B revenues, payments to contract pharmacies, and revenue distribution across the top 50 drug products driving program discounts.^9 10^

Despite these state-level reforms, comprehensive federal transparency requirements remain absent. As a result, the 340B program faces growing scrutiny regarding its accountability, financial stewardship, and alignment with its original mission to serve vulnerable patient populations.^8^

## Objectives

The current study compares reported charity care, unreimbursed or uncompensated care, and Medicaid patient volume among 340B and non-340B hospitals as measures relevant to the 340B program’s policy goal of supporting care for uninsured and underserved patients. These comparisons are intended to serve as a policy accountability benchmark rather than a causal assessment of 340B participation.

## Methods

### Data Source

We used RAND-transposed CMS HCRIS hospital cost report data obtained on May 1, 2025. The analysis relied on the reported values in the RAND-transposed file, and the authors made no additional adjustments to the underlying reported cost report values. 340B designation was derived by linking hospitals in the RAND-transposed CMS HCRIS hospital cost report data to HRSA 340B covered-entity records. Hospitals were classified as 340B participants if they were identified as active 340B covered entities during the relevant reporting period. Hospitals without an active HRSA 340B designation were classified as non-340B.

### Measures

To assess reported financial assistance and patient-volume measures relevant to low-income and uninsured patients, the following measures were evaluated:

- Charity care as a percentage of operating expenses;
- Medicaid admission days as a percentage of total hospital days;
- Unreimbursed or uncompensated care as a percentage of operating expenses;
- Cost of initial obligation of uninsured patients approved for charity care as a percentage of operating expenses; and
- Charity care for uninsured patients as a percentage of operating expenses.

### Hospital Comparisons

We compared mean values for 340B and non-340B hospitals overall and across selected 340B hospital categories and subtypes. Comparisons included:

- All 340B hospitals compared with all non-340B hospitals; and
- 340B hospitals with DSH, CAH, RRC, and SCH designations compared with all non-340B hospitals and with all 340B hospitals.

### Exclusion Criteria

Pediatric hospitals were excluded because, in almost all cases, they did not report charity care data, and including them would have produced inconsistent denominators across the measures used in the analytic comparisons.

To ensure consistent reporting across measures and maintain the integrity of the denominator for each category, any hospital that did not report all of the following parameters was excluded from the final analysis:

- Operating expenses
- Charity care as a percentage of operating expenses
- Medicaid admission days as a percentage of total hospital days
- Medicare admission days as a percentage of total hospital days
- Non-Medicaid/Medicare admission days as a percentage of total hospital days
- Unreimbursed or uncompensated care as a percentage of operating expenses
- Net income
- Non-Medicare bad debt as a percentage of operating expenses
- Cost of initial obligation of uninsured patients approved for charity care as a percentage of operating expenses

## Statistical Analysis

We conducted a cross-sectional analysis comparing unadjusted mean values across 340B and non-340B hospital categories. Because hospital counts differed across 340B and non-340B categories and subtypes, and because equal variances across groups could not be assumed, Welch’s t-test for independent samples with unequal variances was used to assess whether differences in mean reported measures were statistically significant.

Welch’s t-tests were applied to pre-specified group comparisons. These included comparisons of all 340B hospitals with all non-340B hospitals and comparisons of 340B hospital subcategories with both all non-340B hospitals and all 340B hospitals. Statistical significance was assessed at P<.05. The statistical testing evaluated differences in unadjusted mean values and should therefore be interpreted as identifying statistically significant differences between hospital groups, not as estimating causal effects of 340B participation.

## Results

As shown in Table 1, 340B hospitals reported significantly lower mean charity care than non-340B hospitals (2.16% vs 2.82% of operating expenses, P<.05) and significantly lower charity care for uninsured patients (1.60% vs 2.26%, P<.05) as a share of operating expenses. 340B hospitals also reported significantly lower initial obligations of uninsured patients approved for charity care than non-340B hospitals (1.61% vs 2.27%, P<.05). Unreimbursed or uncompensated care was numerically lower among all 340B hospitals than non-340B hospitals (6.90% vs 7.18% of operating expenses), but this overall difference was not statistically significant.

**Table 1:** Comparison of non-340B and 340B hospitals.

|  | Number<br>of<br>hospitals | Charity<br>care as<br>% of OE | Charity care for<br>uninsured as<br>% of OE | Initial obligation of<br>uninsured patients<br>approved for charity<br>care as % of OE | Unreimbursed/<br>uncompensated<br>care as % of OE<sup>c</sup> |
| --- | --- | --- | --- | --- | --- |
| **Non-340B (all)** | **1559** | **2.82** | **2.26** | **2.27** | **7.18** |
| **340B (all)** | **2440** | **2.16<sup>a</sup>** | **1.60<sup>a</sup>** | **1.61<sup>a</sup>** | **6.90** |
| DSH | 1146 | 2.62<sup>b</sup> | 2.09<sup>b</sup> | 2.10<sup>a</sup> | 7.04 |
| CAH | 1060 | 1.69<sup>a b</sup> | 1.09<sup>a b</sup> | 1.11<sup>a b</sup> | 6.92 |
| RRC | 121 | 2.07<sup>a</sup> | 1.51<sup>a</sup> | 1.52<sup>a</sup> | 6.12<sup>a b</sup> |
| SCH | 113 | 2.00<sup>a</sup> | 1.50<sup>a</sup> | 1.50<sup>a</sup> | 6.20<sup>a</sup> |

<sup>a</sup> Statistically significant difference compared with all non-340B hospitals using Welch's t-test; $P<.05$ .
<sup>b</sup> Statistically significant difference compared with all 340B hospitals using Welch's t-test; $P<.05$ .
<sup>c</sup> Includes Medicaid, CHIP, charity care, and bad debt.
Abbreviations: CAH, critical access hospital; DSH, disproportionate share hospital; RRC, rural referral center; SCH, sole community hospital; OE, operating expenses.

Within the 340B hospital category, several statistically significant differences were observed across hospital subtypes. DSH hospitals reported higher mean charity care and charity care for uninsured patients than all 340B hospitals (2.62% vs 2.16% and 2.09% vs 1.60%, respectively; P<.05). However, DSH hospitals reported significantly lower initial obligations of uninsured patients approved for charity care than non-340B hospitals (2.10% vs 2.27%, P<.05). CAH hospitals reported significantly lower charity care, charity care for uninsured patients, and initial obligations of uninsured patients approved for charity care than both non-340B hospitals and all 340B hospitals (P<.05). RRC and SCH hospitals also reported significantly lower charity care, charity care for uninsured patients, and initial obligations for uninsured patients than non-340B hospitals (P<.05). For unreimbursed or uncompensated care, RRC hospitals reported significantly lower values than both non-340B hospitals and all 340B hospitals, while SCH hospitals reported significantly lower values than non-340B hospitals (P<.05).

Table 2 shows that 340B hospitals reported higher average Medicaid admission days than non-340B hospitals (19.69% vs 17.76%, P<.05). However, CAH hospitals reported lower average Medicaid admission days than non-340B hospitals (8.40% vs 17.76%, P<.05) and all 340B hospitals (8.40% vs 19.69%, P<.05). DSH hospitals reported the highest average Medicaid admission days among all hospital types.

**Table 2:** Comparison of non-340B and 340B hospitals.

|  | Number of hospitals | Medicaid admissions as a % share of hospital days |
| --- | --- | --- |
| **Non-340B (all)** | **1559** | 17.76 |
| **340B (all)** | **2440** | **19.69<sup>a</sup>** |
| DSH | 1146 | 29.89<sup>a b</sup> |
| CAH | 1060 | 8.40<sup>a b</sup> |
| RRC | 121 | 19.64<sup>a</sup> |
| SCH | 113 | 22.12<sup>a b</sup> |

<sup>a</sup> Statistically significant difference compared with all non-340B hospitals using Welch's t-test; $P<.05$ .
<sup>b</sup> Statistically significant difference compared with all 340B hospitals using Welch's t-test; $P<.05$ .
Abbreviations: CAH, critical access hospital; DSH, disproportionate share hospital; RRC, rural referral center; SCH, sole community hospital; OE, operating expenses.

## Discussion

Compared with non-340B hospitals, 340B-participating institutions demonstrated mixed performance on measures related to serving uninsured and underserved populations. Although 340B hospitals reported a larger share of Medicaid admission days, indicating greater engagement with low-income patient populations, they reported lower average charity care and charity care for uninsured patients as a percentage of operating expenses than non-340B hospitals. Unreimbursed or uncompensated care was also numerically lower among 340B hospitals, although the overall difference compared with non-340B hospitals was not statistically significant. These findings raise accountability concerns about whether participation in the 340B program is consistently associated with greater reported financial assistance for patients in need, despite the program’s policy goal of supporting uninsured and underserved populations.

Among 340B entities, DSH hospitals reported higher average charity care and Medicaid admission days than other 340B hospital categories. In contrast, CAH hospitals reported lower average charity care and a lower share of Medicaid admission days. These differences suggest that 340B hospital subcategories vary substantially in the extent to which they report charity care, uncompensated care, and Medicaid patient volume. From a policy accountability perspective, these findings raise the question of whether eligibility standards for 340B hospital participation should more consistently reflect low-income patient burden, similar to the DSH-related eligibility framework, which is tied to a hospital’s disproportionate share adjustment percentage and captures components of its low-income inpatient population, including Medicaid-related inpatient days and Medicare SSI-related days.

Although tax status was not directly evaluated in this analysis, differences in tax obligations and public financial benefits between some 340B and non-340B hospitals remain relevant to the broader policy context for evaluating 340B program accountability and reform. Comparisons with non-340B hospitals should therefore be interpreted as a minimum accountability benchmark rather than a complete measure of program performance. Because 340B hospitals generally operate as nonprofit or public institutions and benefit from tax-exempt status in addition to the federal revenue opportunity created by 340B discounts, parity with non-340B hospitals may be an insufficient policy standard. Non-340B hospitals include both tax-exempt nonprofit institutions and for-profit hospitals that may contribute to local, state, and federal governments through tax payments but do not receive 340B-related revenues. Given these structural differences, policymakers may reasonably expect 340B hospitals to report substantially greater levels of charity care, uncompensated care, and support for uninsured and underserved patients than non-340B hospitals.

The use of Welch’s t-tests helps distinguish statistically significant differences from descriptive variation alone. These findings suggest that 340B participation is associated with higher Medicaid patient volume but does not consistently correspond with higher reported direct financial assistance for uninsured patients. Because the analysis is unadjusted and cross-sectional, the findings should be interpreted as accountability-oriented comparisons rather than causal estimates of the effect of 340B participation.

These results support the need for greater transparency, standardized reporting, and clearer accountability expectations to assess whether 340B-related financial benefits are directed toward the populations the program was intended to support.

### Policy Recommendations

Given these unadjusted cross-sectional findings, federal and state policymakers should consider several policy reforms to strengthen transparency, accountability, and alignment between the 340B program and its intended purpose of supporting uninsured and underserved patients.

- Policymakers should consider whether eligibility standards for 340B hospital participation should more consistently reflect low-income patient burden. One policy option would be to evaluate whether elements of the DSH-related eligibility framework, which is tied to a hospital’s disproportionate share adjustment percentage and captures components of its low-income inpatient population, including Medicaid-related inpatient days and Medicare SSI-related days, should be applied more broadly across 340B hospital covered entities.
- Policymakers should consider minimum charity care and patient-assistance accountability standards for 340B hospitals, including benchmarks for overall charity care and charity care for uninsured patients. Such standards could be informed by national averages among non-340B hospitals or by category-specific benchmarks that account for hospital type and patient population.
- Although federal reporting frameworks include definitions and reporting categories for charity care, hospitals retain substantial discretion in how they operationalize charity care through financial assistance policies, eligibility criteria, patient-screening processes, and accounting practices. Policymakers should strengthen standardized reporting, documentation, and enforcement requirements to assess whether 340B savings are being used to expand direct patient assistance or retained as institutional revenue.
- Policymakers should consider requiring 340B hospitals to separately track and report revenue generated through 340B-related transactions and the corresponding use of those funds. A separate accounting or auditable reporting mechanism could improve transparency and allow policymakers and stakeholders to evaluate whether 340B proceeds are directed toward patient care, charity care, and services for underserved populations.
- To further enhance patient transparency, 340B hospitals should provide a standardized disclosure notice explaining their participation in the 340B Drug Pricing Program and providing contact information for the hospital’s financial assistance and charity care department.

## Limitations

We compared reported mean values across hospital categories using unadjusted descriptive comparisons and Welch’s t-tests for selected reported measures, including charity care, charity care for uninsured patients, unreimbursed or uncompensated care, and Medicaid admission days. However, no risk adjustment or multivariable modeling was performed. The analysis also did not control for differences in hospital size, geography, ownership, payer mix, service lines, tax status, or patient clinical complexity. In addition, multiple pairwise comparisons were performed, and statistical findings should be interpreted in the context of the study’s exploratory, accountability-oriented design. As a result, the findings should be interpreted as observed differences between hospital groups rather than causal estimates of the effect of 340B participation.

## Conclusion

This cross-sectional analysis raises accountability concerns about whether 340B hospital participation is consistently associated with greater reported financial assistance for uninsured and underserved patients. Although 340B hospitals reported a significantly higher share of Medicaid admission days, they reported lower average charity care and charity care for uninsured patients than non-340B hospitals. Unreimbursed or uncompensated care was also numerically lower among 340B hospitals, although the overall difference was not statistically significant.

These findings suggest that 340B participation alone does not consistently correspond with higher reported charity care or direct financial assistance on the measures examined. In the broader policy context, the financial advantages associated with 340B participation and tax-exempt hospital status support the need for stronger transparency, standardized reporting, and clearer accountability expectations to ensure that program benefits are directed toward uninsured and underserved patients.

## Data Availability

RAND Corporation data acquired by Pioneer Institute.

## Notes

Financial support and intellectual input for this paper were provided by Cancer*Care*.

### Competing Interest Statement

The authors have declared no competing interest.

### Summary of Updates

We included statistical analysis to complement the study's findings. The Methods, Results, Discussion, and Conclusion sections reflect that change.

